# A systematic review of the validated monogenic causes of human male infertility: 2020 update and a discussion of emerging gene-disease relationships

**DOI:** 10.1101/2021.05.01.21256465

**Authors:** Brendan J. Houston, Antoni Riera-Escamilla, Margot J. Wyrwoll, Albert Salas-Huetos, Miguel J. Xavier, Liina Nagirnaja, Corinna Friedrich, Don F. Conrad, Kenneth I. Aston, Csilla Krausz, Frank Tüttelmann, Moira K. O’Bryan, Joris A. Veltman, Manon S. Oud

**Author notes:** To whom correspondence should be addressed: Brendan J. Houston -, Bio21 institute, 30 Flemington Rd, Parkville VIC 3052, Australia; Manon S. Oud -, Department of Human Genetics (route 848), Geert Grooteplein Zuid 10, 6525 GA, Nijmegen, the Netherlands.

## Abstract

**Background:** Human male infertility has a notable genetic component, including well established diagnoses like Klinefelter syndrome, Y-chromosome microdeletions, and monogenic causes. Approximately 4% of all infertile men are now diagnosed with a genetic cause, but a vast majority (60-70%) remain without a clear diagnosis and are classified as unexplained. This is likely in large part due to a delay in the field adopting next generation sequencing technologies, and the absence of clear statements from leaders in the field as to what constitutes a validated cause of human male infertility (the current paper aims to address this). Fortunately, there has been a significant increase in the number of male infertility next generation sequencing studies. These have revealed a considerable number of novel gene-disease relationships (GDRs), which each require stringent assessment to validate the strength of genotype-phenotype associations. To definitively assess which of these GDRs are clinically relevant, the International Male Infertility Genomics Consortium (IMIGC) has identified the need for a systematic review and a comprehensive overview of known male infertility genes and an assessment of the extent of evidence for reported GDRs.

**Objective and rationale:** In 2019, the first standardised clinical validity assessment of monogenic causes of male infertility was published. Here, we provide a comprehensive update of the subsequent 1.5 years, employing the joint expertise of the IMIGC to systematically evaluate all available evidence (as of July 1st, 2020) for monogenic causes of isolated or syndromic male infertility, endocrine disorders or reproductive system abnormalities affecting the male sex organs. In addition, we systematically assessed the evidence for all previously reported possible monogenic causes of male infertility, using a framework designed for a more appropriate clinical interpretation of disease genes.

**Search methods:** We performed a literature search according to the PRISMA guidelines up until the 1st of July 2020 for publications in English, using search terms related to “male infertility” in combination with the word “genetics” in PubMed. Next, the quality and the extent of all evidence supporting selected genes was assessed using an established and standardised scoring method. We assessed the experimental quality, patient phenotype assessment, and functional evidence based on gene expression, mutant *in vitro* cell and *in vivo* animal model phenotypes. A final score was used to determine the clinical validity of each GDR, as expressed by the following five categories: no evidence, limited, moderate, strong or definitive. Variants were also reclassified according to the ACMG-AMP guidelines and were recorded in spreadsheets for each GDR, which is available at imigc.org.

**Outcomes:** The primary outcome of this review was an overview of all known GDRs for monogenic causes of human male infertility and their clinical validity. We identified a total of 120 genes that were moderately, strongly or definitively linked to 104 infertility phenotypes.

**Wider implications:** Our systematic review summarises and curates all currently available evidence to reveal the strength of GDRs in male infertility. The existing guidelines for genetic testing in male infertility cases are based on studies published 25 years ago, and an update is far past due. The insights generated in the current review will provide the impetus for an update of existing guidelines, will inform novel evidence-based genetic testing strategies used in clinics, and will identify gaps in our knowledge of male infertility genetics. We discuss the relevant international guidelines regarding research related to gene discovery and provide specific recommendations to the field of male infertility.

## Introduction

Male infertility is a common condition, affecting at least 7% of men worldwide, and is often predicted to be largely genetic in origin (Krausz and Riera-Escamilla, 2018). A majority of all human (84%, 16,598; Uhlen et al. 2015) and mouse (90%, 18,037; Schultz et al. 2003) protein-coding genes are expressed in the testis, emphasising that sperm production is a complex process and involves many separate biological pathways. Further, sperm must mature in the epididymis and undergo a final step of activation in the female reproductive tract, termed ‘capacitation’, before they are capable of fertilising an oocyte. By extension, there are many points during sperm development and maturation that could be compromised by the effect of genetic variants. The identification of genes affected by these variants and the assessment of definitive genotype-phenotype correlations, however, remains a challenge.

Advances in next generation sequencing (NGS) have greatly facilitated the unbiased exome-wide (whole exome sequencing [WES]) and genome-wide (whole genome sequencing [WGS]) detection of any genetic variants that may play a role in male infertility (examples Alhathal et al. 2020; Liu et al. 2021; Chen et al. 2020; Coutton et al. 2019; Nagirnaja et al. submitted; Oud et al. submitted). However, the population frequency of these variants is expected to be very low given their negative effect on fertility. For this reason, the analysis of large patient and control cohorts is essential to identify recurrently mutated genes and detect statistical enrichments in patient cohorts. One major problem in the male infertility field is that genetic testing strategies employed in andrology clinics are not standardised, and in some countries/states even the most basic of tests (karyotype and AZF deletion analysis) are still not routinely used.

The male infertility field is currently catching up to other fields/disease types with a strong genetic component, such as intellectual disability, neuromuscular disorders and hereditary hearing impairments (Markitantova and Simirskii, 2020; Chiu et al. 2020; Whatley et al. 2020). The rapid uptake of NGS technologies in the male infertility research field over the past 5-10 years, as well as the development of international consortia to collect and characterise clinical cohorts, is aiding in the transition of findings into clinical practice. To assist in this feedback of knowledge, clear direction is required for the validity of which individual genes to be screened and their relevance to certain types of infertility.

In this article, we provide an updated clinical validity assessment of the monogenic causes of male infertility (Oud et al. 2019), through the systematic analysis of newly published evidence from January 2019-July 2020 for individual gene-disease relationships (GDRs). We have employed the joint expertise of the IMIGC to systematically evaluate/re-evaluate all available evidence for published monogenic causes of isolated or syndromic male infertility, endocrine disorders that impact male fertility and reproductive system abnormalities affecting the male sex organs. This analysis has resulted in the identification of 104 high-probability ‘human male infertility genes’, a 33% increase from the number identified in 2019.

## Methods

### Search strategy and study selection

A literature search was performed as described in Oud et al. (2019) to identify articles reporting on monogenic causes of male infertility or male reproductive system anomalies entered into MEDLINE-Pubmed before the 1^st^ of July 2020. Assessment of whether the articles met the inclusion or exclusion criteria (detailed in Supplementary Table I) was performed by two independent reviewers (BJH and MSO). The present study and the corresponding search protocol were registered with the PROSPERO registry (http://www.crd.york.ac.uk/PROSPERO) as PROSPERO 2021: CRD42021229164.

### Data extraction and assessment

The clinical validity of each identified GDR was scored using a system published by Smith et al. (2017). Scoring was performed by two randomly assigned reviewers (BJH, AR-E, MJW, AS-H, MJX, LN, CF and MSO) using a standardised assessment template to: (i) extract gene names, inheritance patterns, patient phenotypes, method of discovery (sequencing method), (ii) annotate variants, and (iii) assess both functional evidence and clinical data, including the outcome of assisted reproductive technologies and recorded co-morbidities. Expression of genes across human organs was assessed by consulting data available on the Human Protein Atlas website, NCBI’s RNA-seq dataset (Fagerberg et al. 2014), GTEx, and an unpublished human testis single cell RNA-seq library tool (‘https://conradlab.shinyapps.io/HISTA; Mahyari et al. under review). To avoid bias in gene-disease evaluation and any conflicts of interest, reviewers were not allowed to score any GDRs they had published. Scoring was separated into five categories: no evidence (<3 points), limited (3-8 points), moderate (9-12 points), strong (13-15 points) or definitive (>15 points), with of a maximum of 17 points. After independent scoring, the individual scores (for each GDR) for both reviewers were compared and any inconsistencies in scoring (>1 point difference or a difference in the final classification) were settled by the two assigned reviewers. Where this was not possible, the scoring was discussed with all non-conflicted reviewers. GDRs with a moderate or higher classification were deemed as confidently linked to human male infertility and combined scoring sheets are available at http://www.imigc.org/. All results in the study are collated in Supplementary Table A PRISMA checklist is available for this study as shown in Supplementary Table III.

## Results

### Summary of included studies and design

We performed a literature search using terms related to “male infertility” in combination with keywords related to the word “genetics” in MEDLINE-PubMed and used the same inclusion and exclusion criteria as described previously (Oud et al. 2019). A total of 26,250 articles were identified, of which 2,765 new articles were identified since the previous search in 2018 and not covered by the previous review (Figure 1). The total number of publications on genetic causes of male infertility has increased by 13% from 849 over the period 2010-2014 to 963 articles in the period 2015-2019 (Figure 2A). Further, while the final search performed for this review was completed in mid-2020, the total number of 2020 studies was estimated to overtake the 2019 number by 20%. The absolute and relative contribution of publications on proposed monogenic causes of phenotypes related to male infertility is also growing (46% in 2017 vs. 52% in 2020). The relative contribution of publications on association studies has declined by 42% (27% in 2017 vs. 19% in 2020), Figure 2B). The shift away from Sanger sequencing to NGS methods is continuing to take place, and 71% of all sequencing studies used NGS in 2020 (Figure 2C), which constitutes an increase of 90% since 2017. While WGS has started to emerge in the field (recent examples: Bedoni et al. 2016; Wang et al. 2017; Yuan et al. 2020; Arafat et al. 2020), WES is currently still the predominant method over panel sequencing and whole genome sequencing (75% vs. 19% and 6%, respectively). Nevertheless, there are clinical presentations where Sanger/panel sequencing is still being utilised at a high frequency, e.g. 63% of studies on 46,XX and 46,XY DSDs in the period of 2019-2020.

**Figure 1.**
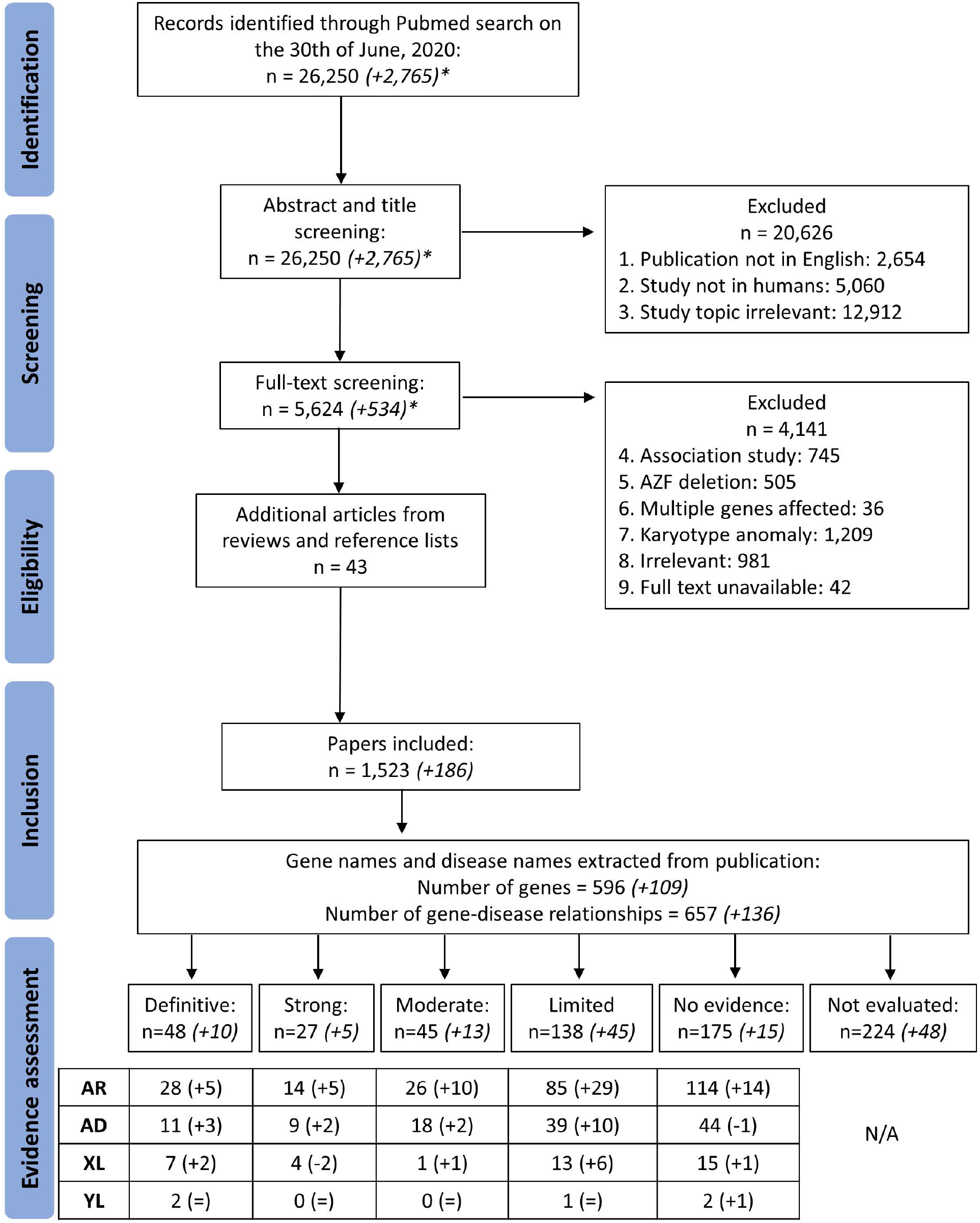
PRISMA Flowchart of search and assessment process.

**Figure 2.**
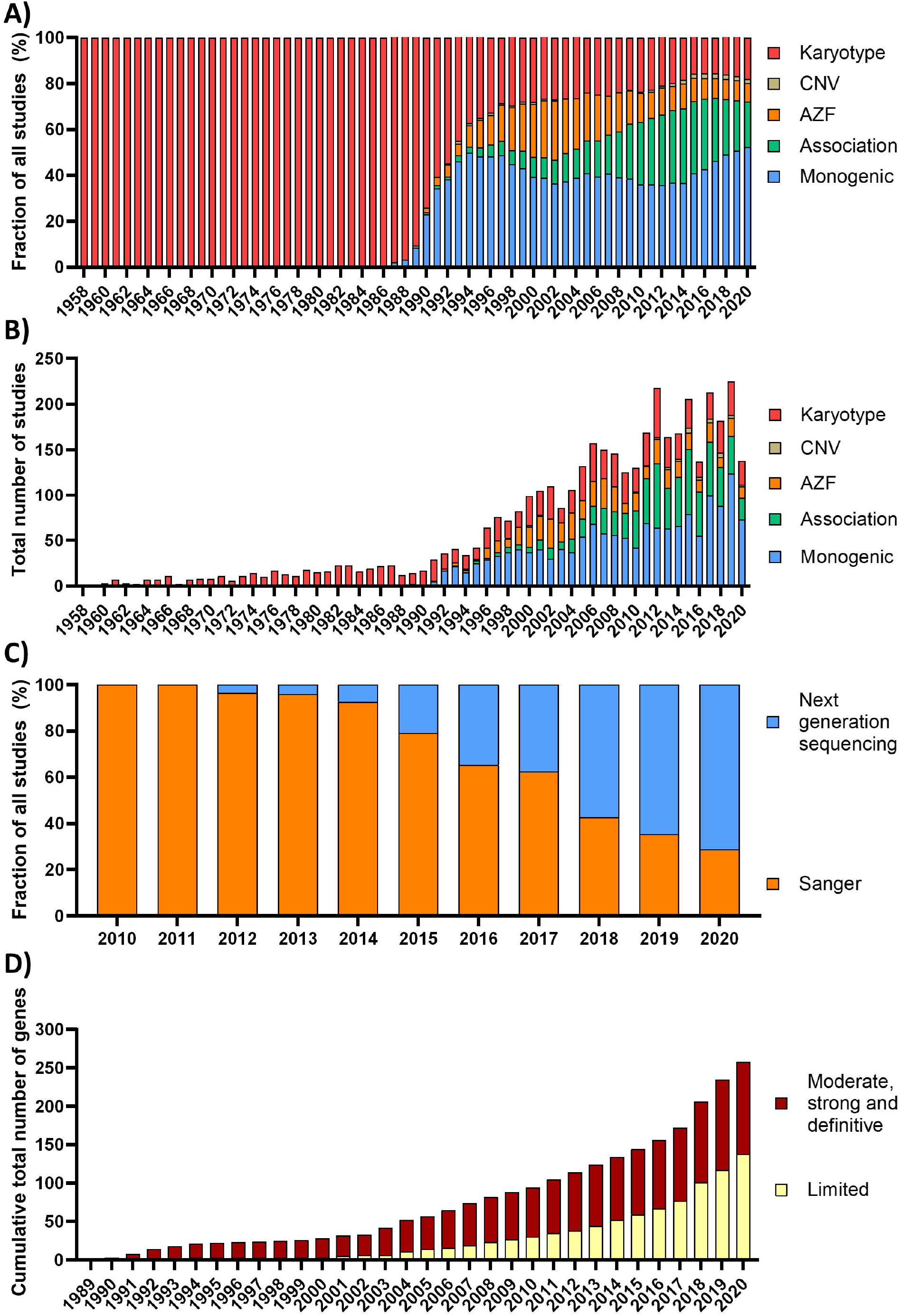
Breakdown of genetic testing approach and number of genes associated with male infertility phenotypes with limited or moderate and higher evidence classification, over time. A. The fraction of studies assessed in this paper that used particular sequencing technologies to elucidate causes of male infertility from 1958-2020, including karyotyping, copy number variation assessment, azoospermia factor region deletion assessment, association studies and studies investigating monogenic causes. B. The total number of studies assessed in this paper investigating male infertility, from 1958-2020, based on sequencing approach C. The fraction of studies assessed in this paper using next generating sequencing technology compared to Sanger sequencing, from 2010-2020. D. The cumulative number of genes and their strength of evidence as linked to male infertility phenotypes based the scoring criteria used in this paper, from 1989-2020.

### Systematic evaluation of evidence

A total of 1,523 publications met all inclusion criteria and were used in this systematic review. Of these, 186 publications were published in the period from 2019 to mid-2020 and were thus not included in the previous clinical validity assessment. Overall, 657 individual GDRs described in these 1,523 publications were investigated, of which 136 were novel and 521 were re-evaluated with our updated assessment criteria in order to incorporate any additional supporting evidence (Supplementary Table II).

The quality and the extent of all evidence for the GDRs was assessed by two independent reviewers, using a standardised scoring method. This assessment was performed for GDRs that were new (n=136), existing with newly published evidence (n=145), or previously confidently linked (moderate, strong or definitive) to male infertility with no newly published evidence (n=19). This score was used to assign a clinical validity of each GDR as: no evidence, limited, moderate, strong, or definitive. We assessed the experimental quality, patient phenotype information, functional evidence based on gene expression and the study of *in vitro* and *in vivo* loss-of-function animal/cell models. Variants were also reclassified according to the widely accepted ACMG-AMP standards and then recorded in spreadsheets for each GDR.

Of the newly identified GDRs, 17 were classified as having moderate evidence or higher (confidently associated with the phenotype) and 43 were classified as having limited evidence. After classification of the existing GDRs that had new evidence published and scoring of GDRs that were previously confidently linked to male infertility with our updated scoring criteria, 45 increased in score since 2019, 11 decreased in score, and 9 changed from unable to be classified to no evidence (or *vice versa*). As detailed above, GDRs were listed as ‘unable to classify’ when the predicted inheritance pattern or genotypes were not reported. In comparison to the previous assessment, 456 scores did not change because no (or insufficient) new evidence was published during the search period (Supplementary Table II).

The novel and established GDRs combined resulted in a total of 104 genes confidently linked to 120 human male infertility or abnormal genitourinary development phenotypes with moderate (n=45), strong (n=27) or definitive (n=48) evidence (Tables I and II). An overview of the organ or cell-level role(s) of these genes is detailed in Figure 3. Another 138 GDRs were classified as ‘limited’ and are thus candidate genes whose dysfunction may result in a male infertility disorder (Supplementary Table II). We propose that latter (limited evidence) group will be of particular interest for review in the next few years.

**Table 1.** Numbers of genes that are at least moderately linked to male infertility or abnormal genitourinary development phenotypes. AR: Autosomal recessive; AD: Autosomal dominant; XL: X-linked; YL: Y-linked

| **Description** | **AR** | **AD** | **XL** | **YL** | **Total** |
| --- | --- | --- | --- | --- | --- |
| **Isolated infertility** | **27** | **5** | **4** | **0** | **36** |
| Acephalic sperm | 3 | 0 | 0 | 0 | 3 |
| Globozoospermia | 1 | 0 | 0 | 0 | 1 |
| Macrozoospermia | 1 | 0 | 0 | 0 | 1 |
| Multiple morphological abnormalities of the sperm flagella | 13 | 1 | 0 | 0 | 14 |
| Non-obstructive azoospermia or oligozoospermia | 7 | 4 | 3 | 0 | 14 |
| Congenital bilateral absence of the vas deferens | 1 | 0 | 1 | 0 | 2 |
| Fertilisation failure | 1 | 0 | 0 | 0 | 1 |
| **Syndromic infertility** | **13** | **3** | **1** | **0** | **17** |
| Primary ciliary dyskinesia | 7 | 0 | 1 | 0 | 8 |
| Other syndromes | 6 | 3 | 0 | 0 | 9 |
| **Endocrine disorder/Reproductive system syndrome** | **28** | **30** | **7** | **2** | **67** |
| Disorders of sexual development | 14 | 13 | 4 | 2 | 33 |
| Hypogonadotropic hypogonadism | 14 | 17 | 3 | 0 | 34 |

**Table 2.** **List of genes linked to male infertility or abnormal genitourinary development phenotypes classified as moderate evidence or higher**.

| Gene | Location | Disorder | Inheritance pattern | Score | Conclusion |
| --- | --- | --- | --- | --- | --- |
| **Isolated infertility** |  |  |  |  |  |
| **ADGRG2** | Xp22.13 | Congenital bilateral absence of the vas deferens; OMIM:300985 | XL | 16 | Definitive |
| **AR** | Xq12 | Non-obstructive azoospermia; OMIM:NA | XL | 17 | Definitive |
| **ARMC2** | 6q21 | Multiple morphological abnormalities of the sperm flagella; OMIM:618433 | AR | 11 | Moderate |
| **AURKC** | 19q13.43 | Macrozoospermia; OMIM:243060 | AR | 17 | Definitive |
| **CFAP251** | 12q24.31 | Multiple morphological abnormalities of the sperm flagella; OMIM:NA (PS258150) | AR | 17 | Definitive |
| **CFAP43** | 10q25.1 | Multiple morphological abnormalities of the sperm flagella; OMIM:617592 | AR | 17 | Definitive |
| **CFAP44** | 3q13.2 | Multiple morphological abnormalities of the sperm flagella; OMIM:617593 | AR | 17 | Definitive |
| **CFAP65** | 2q35 | Multiple morphological abnormalities of the sperm flagella; OMIM:618664 | AR | 15 | Strong |
| **CFAP69** | 7q21.13 | Multiple morphological abnormalities of the sperm flagella; OMIM:617959 | AR | 13 | Strong |
| **CFAP91** | 3q13.33 | Multiple morphological abnormalities of the sperm flagella; OMIM:609910 | AR | 9 | Moderate |
| **CFTR** | 7q31.2 | Congenital bilateral/unilateral absence of vas deferens; OMIM:277180 | AR | 17 | Definitive |
| **DMRT1** | 9p24.3 | Non-obstructive azoospermia; OMIM:NA(PS258150) | AD | 10 | Moderate |
| **DNAH1** | 3p21.1 | Multiple morphological abnormalities of the sperm flagella; OMIM:617576 | AR | 17 | Definitive |
| **DNAH17** | 17q25.3 | Multiple morphological abnormalities of the sperm flagella; OMIM:618643 | AR | 15 | Strong |
| **DPY19L2** | 12q14.2 | Globozoospermia; OMIM:613958 | AR | 16 | Definitive |
| **FANCM** | 14q21.2 | Oligozoospermia; OMIM:NA (PS258150) | AR | 13 | Strong |
| **FSIP2** | 2q32.1 | Multiple morphological abnormalities of the sperm flagella; OMIM:618153 | AR | 12 | Moderate |
| **KLHL10** | 17q21.2 | Oligozoospermia; OMIM:615081 | AD | 10.5 | Moderate |
| **M1AP** | 2p13.1 | Non-obstructive azoospermia; OMIM:619108 | AR | 12 | Moderate |
| **MEI1** | 22q13.2 | Non-obstructive azoospermia; OMIM:NA (PS258150) | AR | 13 | Strong |
| **PLCZ1** | 12p12.3 | Fertilization failure; OMIM:617214 | AR | 16 | Definitive |
| **PMFBP1** | 16q22.2 | Acephalic spermatozoa; OMIM:618112 | AR | 14 | Strong |
| **QRICH2** | 17q25.1 | Multiple morphological abnormalities of the sperm flagella; OMIM:618341 | AR | 12 | Moderate |
| **SEPTIN12** | 16p13.3 | Multiple morphological abnormalities of the sperm flagella; OMIM:614822 | AD | 11.5 | Moderate |
| **SPEF2** | 5p13.2 | Multiple morphological abnormalities of the sperm flagella; OMIM:618751 | AR | 14.75 | Strong |
| **STAG3** | 7q22.1 | Non-obstructive azoospermia; OMIM:NA (PS258150) | AR | 11.5 | Moderate |
| **SUN5** | 20q11.21 | Acephalic sperm; OMIM:617187 | AR | 16.75 | Definitive |
| **SYCP2** | 20q13.33 | Severe oligozoospermia; OMIM:258150 | AD | 10.75 | Moderate |
| **SYCP3** | 12q23.2 | Non-obstructive azoospermia; OMIM:270960 | AD | 14 | Strong |
| **TEX11** | Xp11 | Non-obstructive azoospermia; OMIM:309120 | XL | 16 | Definitive |
| **TEX14** | 17q22 | Non-obstructive azoospermia; OMIM:617707 | AR | 10.5 | Moderate |
| **TEX15** | 8p12 | Non-obstructive azoospermia; OMIM:617960 | AR | 13.5 | Strong |
| **TSGA10** | 2q11.2 | Acephalic spermatozoa; OMIM:617961 | AR | 10.25 | Moderate |
| **TTC29** | 4q31.22 | Multiple morphological abnormalities of the sperm flagella; OMIM:618745 | AR | 14.5 | Strong |
| **USP26** | Xq26.2 | Azoospermia or oligozoospermia; OMIM:NA (PS258150) | XL | 9.5 | Moderate |
| **XRCC2** | 7q36.1 | Non-obstructive azoospermia; OMIM: 617247 | AR | 10 | Moderate |
| **Syndromic infertility** |  |  |  |  |  |
| **APOA1** | 11q23.3 | Testicular amyloidosis; OMIM:105200 | AD | 12 | Moderate |
| **CATSPER2** | 15q15.3 | Deafness infertility syndrome; OMIM: 611102 | AR | 11 | Moderate |
| **CCDC39** | 3q26.33 | Primary ciliary dyskinesia; OMIM:613807 | AR | 13 | Strong |
| **CCDC40** | 17q25.3 | Primary ciliary dyskinesia; OMIM:613808 | AR | 13.25 | Strong |
| **CDC14A** | 1p21.2 | Oligoasthenoteratozoospermia OMIM:608653 | AR | 9 | Moderate |
| **CEP290** | 12q21.32 | Leber congenital amaurosis; OMIM:611755 | AR | 9 | Moderate |
| **DNAAF2** | 14q21.3 | Primary ciliary dyskinesia; OMIM:612518 | AR | 12.25 | Moderate |
| **DNAAF4** | 15q21.3 | Primary ciliary dyskinesia; OMIM:615482 | AR | 13 | Strong |
| **DNAAF6** | Xq22.3 | Primary ciliary dyskinesia: OMIM:300991 | XL | 15 | Strong |
| **FANCA** | 16q24.3 | Occult Fanconi anemia; OMIM:NA (PS227650) | AR | 10 | Moderate |
| **LRRC6** | 8q24.22 | Primary ciliary dyskinesia; OMIM:614935 | AR | 13.5 | Strong |
| **MNS1** | 15q21.3 | Asthenoteratozoospermia; OMIM:NA (PS258150) | AR | 9.5 | Moderate |
| **NLRP3** | 1q44 | Muckle-Wells Syndrome; OMIM:191900 | AD | 9 | Moderate |
| **PKD1** | 16p13.3 | Polycystic kidney disease and asthenozoospermia; OMIM:173900 | AD | 11.25 | Moderate |
| **RSPH3** | 6q25.3 | Primary ciliary dyskinesia; OMIM:616481 | AR | 10.25 | Moderate |
| **SPEF2** | 5p13.2 | Primary ciliary dyskinesia with multiple morphological abnormalities of the sperm flagellum; OMIM:618751 | AR | 12 | Moderate |
| **TRIM37** | 17q22 | Mulibrey nanism; OMIM:253250 | AR | 10 | Moderate |
| **Reproductive system syndrome/endocrine disorder** |  |  |  |  |  |
| **AMH** | 19p13.3 | Persistent Müllerian duct syndrome; OMIM:261550 | AR | 17 | Definitive |
| **AMHR2** | 12q13.13 | Persistent Müllerian duct syndrome; OMIM:261550 | AR | 17 | Definitive |
| **ANOS1** | Xp22.31 | Kallmann syndrome; OMIM:308700 | XL | 16 | Definitive |
| **ANOS1** | Xp22.31 | Isolated hypogonadotropic hypogonadism (normosmic); OMIM:308700 | XL | 13 | Strong |
| **AR** | Xq12 | Partial androgen insensitivity syndrome; OMIM:312300/300633 | XL | 17 | Definitive |
| **BMP4** | 14q22.2 | Hypospadias; OMIM:NA (PS300633). Micropenis; OMIM:NA | AD | 10.25 | Moderate |
| **BMP7** | 20q13.31 | Hypospadias; OMIM:NA (PS300633) | AD | 10.25 | Moderate |
| **BNC2** | 9p22.3-p22.2 | Hypospadias; OMIM:NA (PS300633) | AD | 10 | Moderate |
| **CCDC141** | 2q31.2 | Kallmann syndrome; OMIM:NA (PS147950) | AR | 12 | Moderate |
| **CHD7** | 8q12.2 | Kallmann syndrome without CHARGE phenotype; OMIM:612370 | AD | 16 | Definitive |
| **CHD7** | 8q12.2 | Isolated hypogonadotropic hypogonadism (normosmic) without CHARGE phenotype; OMIM:612370 | AD | 17 | Definitive |
| **CYP11A1** | 15q24.1 | Congenital adrenal insufficiency with partial 46,XY sex reversal (Prader stage 4; 5 or 6); OMIM:613743 | AR | 16 | Definitive |
| **CYP11B1** | 8q24.3 | 46,XX Disorders of sexual development (Prader scale 4; 5 or 6) due to congenital adrenal hyperplasia (11-beta-hydroxylase deficiency); OMIM: 202010 | AR | 17 | Definitive |
| **CYP17A1** | 10q24.32 | 46,XY Disorders of sexual development (Prader stage 4; 5 or 6) due to 17-alpha-hydroxylase/17,20-lyase deficiency; OMIM:202110 | AR | 16 | Definitive |
| **CYP19A1** | 15q21.2 | Aromatase excess syndrome with gynaecomastia; OMIM:139300 | AD | 17 | Definitive |
| **CYP19A1** | 15q21.2 | 46,XX Disorders of sexual development (Prader scale 4; 5 or 6) due to aromatase deficiency; OMIM:613546 | AR | 16 | Definitive |
| **CYP19A1** | 15q21.2 | Male infertility in 46,XY men due to aromatase deficiency; OMIM:613546 | AR | 9.5 | Moderate |
| **CYP21A2** | 6p21.33 | Classic congenital adrenal hyperplasia; OMIM:201910 | AR | 17 | Definitive |
| **CYP21A2** | 6p21.33 | Non-classic adrenal hyperplasia (late onset or no CAH symptoms); OMIM: 201910 | AR | 17 | Definitive |
| **DHX37** | 12q24.31 | 46,XY Disorders of sexual development (Prader scale 4; 5 or 6); OMIM:273250 | AD | 11 | Moderate |
| **FGF17** | 8p21.3 | Kallmann syndrome; OMIM:615270 | AD | 9 | Moderate |
| **FGF8** | 10q24.32 | Kallmann syndrome; OMIM: 612702 | AD | 10 | Moderate |
| **FGF8** | 10q24.32 | Isolated hypogonadotropic hypogonadism (normosmic); OMIM:612702 | AD | 14 | Strong |
| **FGFR1** | 8p11.23 | Kallmann syndrome; OMIM:147950 | AD | 17 | Definitive |
| **FGFR1** | 8p11.23 | Isolated hypogonadotropic hypogonadism (normosmic); OMIM:147950 | AD | 17 | Definitive |
| **FSHB** | 11p14.1 | Isolated hypogonadotropic hypogonadism; OMIM:229070 | AR | 12.25 | Moderate |
| **FSHR** | 2p16.3 | Hypergonadotropic hypogonadism; OMIM:NA (PS147950) | AR | 11 | Moderate |
| **GATA4** | 8p23.1 | 46,XY Disorders of sexual development (Prader scale 4; 5 or 6) resulting in anomalies of testicular development; OMIM:615542 | AD | 13 | Strong |
| **GNRH1** | 8p21.2 | Isolated hypogonadotropic hypogonadism; OMIM:614841 | AR | 13.5 | Strong |
| **GNRHR** | 4q13.2 | Isolated hypogonadotropic hypogonadism; OMIM:146110 | AR | 17 | Definitive |
| **HS6ST1** | 2q14.3 | Kallmann syndrome; OMIM:614880 | AD | 9.5 | Moderate |
| **HSD17B3** | 9q22.32 | 46,XY Disorders of sexual development (Prader scale 4; 5 or 6) resulting in anomalies of testicular development; OMIM:264300 | AR | 16 | Definitive |
| **HSD3B2** | 1p12 | Adrenal hyperplasia due to 3β-hydroxysteroid dehydrogenase deficiency; OMIM:201810 | AR | 16.5 | Definitive |
| **IGSF10** | 3q25.1 | Delayed puberty; OMIM:NA (PS147950) | AD | 9.25 | Moderate |
| **IL17RD** | 3p14.3 | Kallmann syndrome with hearing loss; OMIM:615267 | AD | 14.5 | Strong |
| **INSL3** | 19p13.11 | Cryptorchidism; OMIM:219050 | AD | 12 | Moderate |
| **KISS1R** | 19p13.3 | Kallmann syndrome; OMIM:614837 | AR | 9 | Moderate |
| **KISS1R** | 19p13.3 | Isolated hypogonadotropic hypogonadism (normosmic); OMIM:614837 | AR | 17 | Definitive |
| **LHB** | 19q13.33 | Isolated Hypogonadotropic hypogonadism; OMIM:228300 | AR | 16.5 | Definitive |
| **LHCGR** | 2p16.3 | Leydig cell dysfunction with hypogonadism; OMIM:238320 | AR | 16.5 | Definitive |
| **LHCGR** | 2p16.3 | Male precocious puberty; OMIM:176410 | AD | 17 | Definitive |
| **MAMLD1** | Xq28 | 46,XY Disorders of Sex Development (Prader scale 4; 5 or 6); OMIM:300758 | XL | 15 | Strong |
| **MYRF** | 11q12.2 | 46XY Disorders of Sex Development, OMIM gene 608329 | AD | 14.5 | Strong |
| **NR0B1** | Xp21.2 | Congenital adrenal hypoplasia; OMIM:300200 | XL | 17 | Definitive |
| **NR0B1** | Xp21.2 | Late-onset adrenal failure or isolated hypogonadotropic hypogonadism; OMIM:NA (PS147950) | XL | 17 | Definitive |
| **NR5A1** | 9q33.3 | 46,XY Disorders of sexual development (Prader scale 4; 5 or 6); OMIM:612965 | AD | 17 | Definitive |
| **NR5A1** | 9q33.3 | 46,XX Disorders of sexual development (Prader scale 4; 5 or 6); OMIM:617480 | AD | 16 | Definitive |
| **NR5A1** | 9q33.3 | Isolated spermatogenic failure; OMIM:184757 | AD | 14 | Strong |
| **PLXNA1** | 3q21.3 | Kallmann syndrome; OMIM:NA (PS147950) | AD | 13.5 | Strong |
| **POU1F1** | 3p11.2 | Combined pituitary hormone deficiency; OMIM:613038 | AR | 16 | Definitive |
| **PROK2** | 3p13 | Kallmann syndrome; OMIM:610628 | AR | 11.5 | Moderate |
| **PROKR2** | 20p12.3 | Kallmann syndrome; OMIM:244200 | AR | 17 | Definitive |
| **PROP1** | 5q35.3 | Pituitary hormone deficiency; OMIM:262600 | AR | 17 | Definitive |
| **RSPO1** | 1p34.3 | Palmoplantar hyperkeratosis with squamous cell carcinoma of skin and sex reversal;<br>OMIM:610644 | AR | 12 | Moderate |
| **SEMA3A** | 7q21.11 | Kallmann syndrome; OMIM:614897 | AD | 16 | Definitive |
| **SOX10** | 22q13.1 | Kallmann syndrome; OMIM:NA (PS147950) | AD | 16 | Definitive |
| **SOX2** | 3q26.33 | Isolated hypogonadotropic hypogonadism (normosmic); OMIM:NA (PS147950) | AD | 16 | Definitive |
| **SOX3** | Xq27.1 | 46,XX Disorders of sexual development (Prader scale 4; 5 or 6); OMIM:NA | XL | 13 | Strong |
| **SOX9** | 17q24.3 | 46,XY Disorders of sexual development (Prader scale 4; 5 or 6); OMIM:NA | AD | 13.5 | Strong |
| **SRD5A2** | 2p23.1 | 46,XY Disorders of sexual development (Prader scale 4; 5 or 6); OMIM:264600 | AR | 17 | Definitive |
| **SRY** | Yp11.2 | 46,XX Disorders of sexual development (Prader scale 4; 5 or 6); OMIM:400045 | YL | 17 | Definitive |
| **SRY** | Yp11.2 | 46,XY Disorders of sexual development (Prader scale 4; 5 or 6); OMIM:400044 | YL | 17 | Definitive |
| **STAR** | 8p11.23 | Lipoid adrenal hyperplasia; OMIM:201710 | AR | 10 | Moderate |
| **TACR3** | 4q24 | Kallmann syndrome; OMIM:614840 | AR | 16.5 | Definitive |
| **WDR11** | 10q26.12 | Hypogonadotropic hypogonadism; OMIM:614858 | AD | 12 | Moderate |
| **WDR11** | 10q26.12 | Kallmann syndrome; OMIM:614858 | AD | 11 | Moderate |
| **WT1** | 11p13 | 46,XY Disorders of sexual development (Prader scale 4; 5 or 6) without Wilm's tumor;<br>OMIM:NA (PS400044) | AD | 14.25 | Strong |

**Figure 3.**
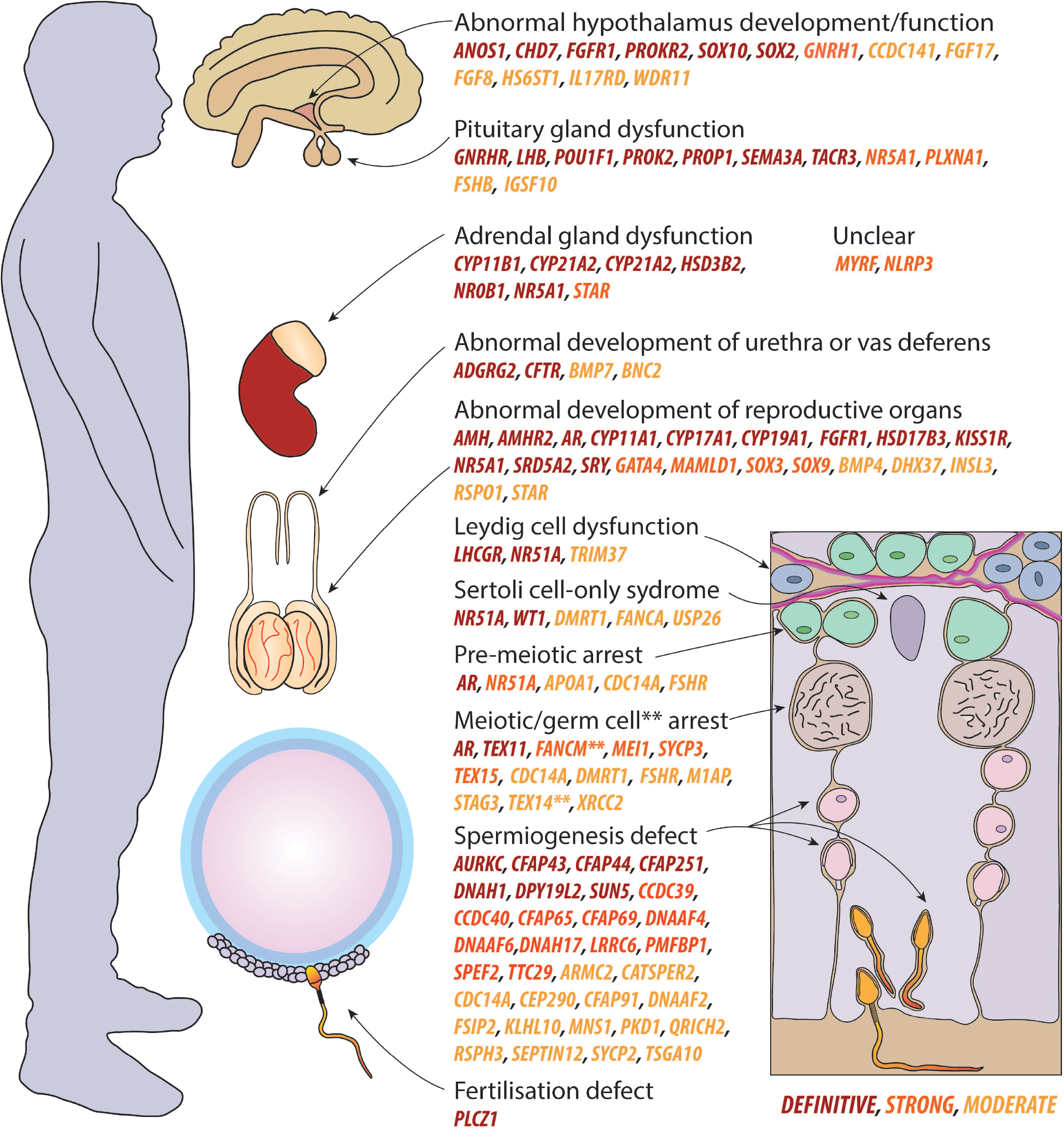
Overview of all genes associated with male infertility phenotypes at an organ/cell level. Definitive genes are labelled in red, strong in orange and moderate in yellow. Organs top to bottom: brain, adrenal gland (and kidney), testes and epididymides with vas deferens. Bottom left: sperm fertilising an oocyte surrounded by cumulus cells. Right: seminiferous tubule cross-section. Leydig cells (blue), Sertoli cells (purple), basement membrane (pink), spermatogonia (green), spermatocytes (brown), spermatids (pink and orange). ** denotes germ cell arrest gene for *FANCM* and *TEX14*. Genes were classified as unclear when they were not clearly linked to a specific organ.

### Insights into the genetic causes of male infertility

As outlined above, in recent years, the identification of genes confidently linked to a male infertility phenotype has risen by 150% from a stable average of approximately 4 per year in the period of 2000-2016 to 10 per year in the period of 2017-2019 (Figure 2C). Additionally, the number of GDRs with limited (emerging) evidence is growing quickly, at approximately 17 new genes per year in the period 2017-2019 (Figure 2C).

The majority of confident GDRs were endocrine disorders or reproductive system syndromes (n=67) and isolated infertility phenotypes (n=36), while a minority were linked to syndromic infertility (n=17), including primary ciliary dyskinesia (Table I). The systematic analysis of patients who present with multiple morphological abnormalities of the sperm flagella (MMAF) phenotypes was also of particular note. In 2018 there were 5 genes confidently linked to human MMAF. This has now tripled to a total of 15 genes as of mid-2020. Although representing only a small fraction of patients with isolated infertility, the use of NGS has the potential to diagnose up to ∼50% of MMAF patients (Toure et al. 2021). A further 15 genes were confidently linked to the most frequently presenting clinical presentations, NOA and oligozoospermia (Tables I and II). The majority of all confident GDRs represented an autosomal recessive inheritance pattern (n=68), while autosomal dominant (n=38), X-linked (n=12) and Y-linked (n=2) inheritance patterns were also reported.

## Discussion

The aim of this study was to provide an updated assessment of all genetic variants reported as causative of human male infertility phenotypes. We also highlight male infertility phenotypes that have received significant attention in the last few years and identify a group of promising new candidate genes. These genes are currently classified as limited evidence, requiring additional replication studies and/or functional evidence in order to be classified as strongly associated with male infertility. Based on our findings, the authors/the IMIGC consortium recommend several updates to the genetic testing standards currently employed in the field of human male infertility, most importantly being the adoption of exome sequencing as the default sequencing approach.

Our clinical validity assessment revealed that as of the 1st of July 2020, a total of 104 genes were linked to a total of 120 male infertility or abnormal genitourinary development phenotypes. This is a significant increase from the 2019 report wherein 78 genes (33% increase to 104) were associated with 92 phenotypes (30% increase to 120) at a moderate or higher level of supporting evidence. As this previous assessment included all published reports from 1958 to 2018, this significant increase in two years further emphasises the healthy uptake of studies to elucidate genetic causes of male infertility, specifically those using NGS.

### Recent developments in genetics of male infertility research and diagnostics

The diagnostic rate of genetic tests for all types of isolated male infertility combined currently sits between 4 and 9.2% (Tüttelmann et al. 2018; Punab et al. 2017; Olesen et al. 2017). These rates are notably behind levels seen in other heterogeneous disorders with a large genetic contribution such as developmental delay (∼30%) and cardiomyopathies (30-40%) (Rehm, 2017). The biggest difference between these diseases is the slow uptake of NGS approaches in the male infertility field in research, but even more so in diagnostics. WES and WGS are now routinely being applied in the diagnostic follow-up of patients with other genetic disorders, resulting in the availability of very large cohorts for disease gene discovery (e.g. Kaplanis et al. 2020, Bourinaris et al. 2020, Cuvertino et al. 2020). This, unfortunately, is still not the case for severe forms of male infertility in most countries. However, as highlighted in Figures 2C/D, the field of male infertility genetics is now expanding rapidly, largely due to the reduced costs and increasing accessibility of NGS, which allows for a more complete and economic testing of patients. Consequently, the number of novel candidate genes and validated disease genes is rapidly growing, which is essential for further diagnostic implementation of these approaches in male infertility.

#### Multiple morphological abnormalities of the sperm flagellum

The phenotype where most progress has been made in recent years is MMAF. Of the 31 newly discovered genes confidently linked to male infertility, 9 were linked to MMAF (29%). When just considering genes likely associated with primary male infertility (i.e. spermatogenesis genes), this constitutes 47% of new, confidently classified genes (9 of 19). Currently, a total of 14 genes (*ARMC2, CFAP43, CFAP44, CFAP65, CFAP69, CFAP91, CFAP251, DNAH1, DNAH17, FSIP2, QRICH2, SEPTIN12, SPEF2* and *TTC29*) are confidently linked to MMAF and another 9 (*AK7, AKAP4, CEP135, CFAP70, DNAH2, DNAH6, DZIP1, TTC21A* and *WDR19*) are listed as candidate genes, i.e., they have a limited classification. As a reflection of collaborative research efforts, MMAF associated genes also comprise 42% of all confident genes that function during the spermiogenesis period.

Further, and in line with the highly conserved core structure of motile cilia and flagella across tissues, there is a clear phenotypic continuum of patients with phenotypes ranging from classical primary ciliary dyskinesia (PCD), manifesting as complete sperm immotility but normal cytology, to severe forms of teratozoospermia. Sperm tail development is a remarkable process that requires the expression of more than 1,000 proteins (Toure et al. 2021) and their coordinated transport into a distinct ciliary compartment originating from a modified centriole that docks to the sperm head (Pleuger et al. 2020). Thus, there are likely many additional genes required for human/mammalian sperm tail development to be discovered. Novel evidence has been identified from MMAF studies, where variants in *SPEF2* cause PCD with MMAF (Tu et al. 2020). While the origin of this commonality is largely unexplored, it may reflect shared protein transport pathways into the ciliary/sperm tail compartment (Pleuger et al. 2020). There are also genes that play important roles in axoneme function (e.g. *DNAH17* Whitfield et al. 2019), whose loss of function results in an isolated infertility phenotype where cilia are unaffected.

#### Azoospermia and oligozoospermia

NOA and severe oligozoospermia are expected to have extensive genetic heterogeneity due to the multiple phases of spermatogenesis that can be affected to cause these presentations. Although a few genes confidently linked to primary testicular failure are beginning to emerge, including *M1AP, STAG3, SYCP2* and *TEX11*, where data from animal models indicate each is essential for meiosis (Arango et al. 2013; Winters et al. 2014; Yang et al. 2006; Yatsenko et al. 2015), large cohort sizes are critical to reveal the full spectrum of disease genes. The discovery and validation of three of these meiosis genes has been possible through the collaborative efforts of the IMIGC/GEMINI consortia and the use of large infertile cohorts and replication studies. This has now led to the availability of WES data for >3,000 men with NOA or severe oligozoospermia, which underscores the importance of data sharing/collaboration for the continued identification of novel, rare genetic causes of male infertility. In addition, these large cohorts help to validate and better estimate the role of previously implicated variants/genes. For example, in 2017 variants in the D-box region of *PIWIL1* were implicated as a recurrent cause of NOA (Gou et al. 2017). Using WES on 2,740 men with NOA or severe oligozoospermia, the IMIGC/GEMINI consortia demonstrated that pathogenic variants in the D-box region of *PIWIL1* specifically and variants elsewhere in the gene, are not a common cause of male infertility (Oud et al. 2021).

### Identifying the variant(s) causative for male infertility

Currently, approximately 33% of all identified GDRs proposed in the literature fell into the “unable to classify” category. For most, this classification is given because the inheritance pattern is unclear. The list, however, likely contains multiple genes that play an important role in the aetiology of monogenic male infertility. As with any disease, but perhaps exacerbated with the wide range of individual biological processes required for male fertility, it can be difficult to determine the role of poorly characterised genes, and whether loss of their function results in a disease state. To assist in filtering for high confidence variants, members of the GEMINI consortium have developed the ‘Population Sampling Probability’ pipeline (PSAP; Wilfert et al. 2016). This is particularly useful when it is the first time a damaging variant has been found in a novel gene, where calculations are based on inheritance model (autosomal dominant, recessive, etc.) and allele frequency. As has been highlighted by the GEMINI consortium (Nagirnaja et al. submitted), this tool is useful for discovering promising new candidate disease genes and individual pathogenic variants, but independent replication and functional validation is still critically important before genes can be used in a diagnostic setting as a ‘validated disease gene’.

### Dynamics of genotype-phenotype mapping

Recent genetic male infertility studies suggest that a high percentage of diagnoses can be revealed, especially in cohorts of men with maturation arrest. A clear determination of the point of arrest (i.e., at spermatogonia or spermatocytes) can assist in refining possibly relevant genes containing variants, based on any known biological function(s). However, very little so far has been done outside of meiotic and spermatogonial arrest. Various genes have also been linked to multiple phenotypic outcomes; thus, the establishment of a clear genotype-phenotype correlation is not necessarily straightforward. One such example is the *TEX14* gene, where pathogenic variation can lead to a Sertoli cell-only phenotype or spermatocyte arrest (Fakhro et al. 2018). The progressive loss of germ cells may occur as a factor of age due to a role in spermatogonial stem cell renewal, as suggested for *Nxf2* knockout mice (Pan et al. 2009). Alternatively, environmental exposures may interact with the genotype to lead to an exacerbation of the phenotype. Variability in the genotype-phenotype relationship has also been identified for single gene variants in reproductive system syndromes/endocrine disorders (Domenice et al. 2016; Goncalves et al. 2017) in 12 of the 13 (92%) confident genes – *ANOS1, AR, CHD7, CYP19A1, CYP21A2, FGF8, FGFR1, KISS1R, LHCGR, NR0B1, NR5A1* and *WDR11*. These disorders are largely complex in that they effect multiple hormonal pathways and tissues beyond the testis.

Other presentations with variability in genotype and phenotype include congenital hypogonadotropic hypogonadism/Kallmann syndrome. In addition to Mendelian causes, incomplete penetrance and oligogenic forms are thought to be causative for these disorders (Maione et al. 2018). One example is the *PROK2* gene, for which autosomal dominant, recessive and oligogenicity have been proposed to be causative (Leroy et al. 2008). For the disorders/differences of sexual development (DSD) spectrum and hypogonadotropic hypogonadism (HH), research groups are now actively exploring the role of oligogenic inheritance in contributing to the disease (Cangiano et al. 2021). For DSDs, this genetic explanation has been proposed following the finding that many patients present with at least 1 variant in 2 individual known or novel DSD genes, particularly when *MAMLD1* is affected (Camats et al. 2020, Li et al. 2020). Similarly, variants in *FGFR1* and other HH genes have shown to cause Kallman syndrome in an oligogenic manner (reviewed in Cangiano et al. 2021).

### Recommendations for genetic testing in male infertility

Although NGS methods are frequently used in research laboratories to study the genetics of male infertility, they have not yet been extensively employed in clinical genetic diagnostics for this condition. While there are no standardised international guidelines for clinical genetic testing in male infertility, a small number of guidelines have been published by professional andrology groups. Examples include guidelines for oligoasthenoteratozoospermia (OAT; Colpi et al. 2018) and Klinefelter syndrome (Zitzmann et al. 2021) by the European Academy of Andrology, and diagnostics/treatment prior to use of assisted reproductive technologies (Toth et al. 2019) by the German Association of the Scientific Medical Societies. However, sperm count cut-offs and genetic testing approaches vary widely between countries. Patients with oligozoospermia, <5 million sperm/ml (EU/US), or azoospermia are generally offered karyotyping and Y-chromosome microdeletion analysis, while *CFTR* testing is recommended for men with suspected congenital bilateral absence of the vas deferens (Krausz and Riera-Escamilla, 2018). These guidelines, or in some cases purely expert opinions, thus limit the genetic testing to the most common infertility presentations and exclude patients with other sperm phenotypes such as teratozoospermia/asthenoteratozoospermia (e.g. globozoospermia and MMAF) and other sperm motility disorders (asthenozoospermia/OAT). According to the average clinical presentation of German men over the last 30 years, this means that at least 40% of infertile men are not being offered any form of genetic testing (Tüttelmann et al. 2018).

In line with the definitive evidence that pathogenic variants in at least 36 genes may result in isolated infertility, we advocate for an update of genetic testing guidelines for male infertility. Ideally, the blood of infertile men should be used for WES to identify any protein-coding variants in a non-biased manner. We appreciate there will often be an issue with a lack of expertise in this technology, or access to sequencing platforms. IMIGC members (imigc.org) are situated across the US, Europe and Australia, and are interested in forming additional collaborations for clinical and research purposes. As a less impactful, backup approach, the genetic targets of panels used in targeting sequencing should at least be updated to include the genes validated in this study (those moderately linked and stronger). Additionally, a wider subset of patients/phenotypes should be sequenced. Such approaches will help to achieve more diagnoses, better personalised treatment, improved risk assessment for the transmission of infertility to offspring, and better counselling for potential health risks in the infertile man. It will also help to improve counselling of azoospermic men prior to testicular biopsy, as finding a genetic diagnosis may help in predicting the chances for successful testicular sperm extraction, which is an incredibly invasive procedure.

### Wider implications of male infertility genetics

Precise phenotyping of men with pathogenic variants will assist in evaluating potential future health risks. Mounting evidence suggests that male infertility may pose as a predictive condition for life-threatening comorbidities in later life, such as various types of cancer and cardiovascular disease (Jensen et al. 2009; Eisenberg et al. 2014; Ehrlich, 2015). While the precise mechanism(s) underpinning this observation are unknown, it is notable that many genes that are highly expressed in the testis and lead to male infertility when mutated are also expressed in other organs, but at considerably lower levels. For example, multiple DNA repair genes linked to infertility and various cancers (reviewed in Nagirnaja et al. 2018), and *TEX11*, which is expressed in the testis and the pancreas according to RNA-seq data (GTEx Portal). As has been made clear above, this is the case for several cilia genes, where men often present with chronic bronchitis and other lung conditions (reviewed in Leigh et al. 2009; Sironen et al. 2020). Thus, additional studies where the health of infertile men is broadly investigated are urgently needed.

Although the topic could constitute an entire article itself, the rapidly increasing use of assisted reproductive technologies worldwide (de Mouzon et al. 2020) is resulting in the propagation of infertility-causing variants throughout the population. As we identify these variants in men who undergo assisted reproductive technologies, we will also be faced with delivering this information back to the patients through genetic counselling services. This is especially relevant for men with Y-linked variants, where any sons will presumably be inflicted with the same infertility phenotype as their affected fathers and thus also require ARTs if wanting their own biological child.

### Strengths and limitations

We used the joint expertise of the IMIGC to systematically evaluate all available evidence (as of July 1st, 2020) for monogenic causes of isolated or syndromic male infertility, endocrine disorders or reproductive system abnormalities affecting the male sex organs. No other published reviews in this field come close to including the approximately 600 genes comprehensively evaluated here. We improved the quality of evidence scoring by including a larger number of reviewers from independent research groups with broad expertise in male infertility. As all research groups have been involved in disease gene discovery of various genes described in this study, we prevented bias as much as possible by not allowing scoring by reviewers who have worked on respective genes.

## Conclusion

Here, we describe an updated clinical validity assessment for a total of 657 male infertility or abnormal genitourinary development GDRs, involving 596 genes. We identified 104 genes linked to 120 phenotypes with sufficient evidence for use in gene panels. These results may help to improve genetic testing in male infertility research and/or diagnostics.

## Supporting information

Supplementary Table III

## Data Availability

All data are available upon request at

## Authors’ roles

BJH, AR-E, MJW, AS-H, MJX, LN, CF, MSO, MKOB and JAV designed and supervised the study; and DFC, KIA, CK, FT assisted in supervising this study. BJH and MSO selected studies for the inclusion and evaluated their quality. The clinical validity assessments were done by BJH, AR-E, MJW, AS-H, MJX, LN, CF and MSO, which were coordinated by BJH and MSO. BJH and MSO wrote the first draft of the manuscript. All authors contributed to the writing and revision process.

## Data availability

The present study and the corresponding search protocol were registered with the PROSPERO registry (http://www.crd.york.ac.uk/PROSPERO) as PROSPERO 2021: CRD42021229164. All scoring spreadsheets are accessible via the IMIGC website (www.imigc.org).

## Funding

National Health and Medical Research Council APP1120356 to MKOB, JAV, DFC and KIA

The Netherlands Organisation for Scientific Research (918-15-667) to JAV as well as an Investigator Award in Science from the Wellcome Trust (209451) to JAV

German Research Foundation sponsored Clinical Research Unit ‘Male Germ Cells: from Genes to Function’ (DFG, CRU326) to CF and FT

National Institutes of Health: Genomics of Spermatogenic Impairment (R01HD078641) to DFC and KIA

Spanish Ministry of Health Instituto Carlos III-FIS-FEDER (Grant number: PI17/01822 PI20/01562) to AR-E and CK

## Acknowledgements

We appreciate the feedback from Raul Piña-Aguilar for his feedback during initial discussion and planning of this study.

## Table legends

**Supplementary Table I. Search strategy, inclusion and exclusion criteria**

**Supplementary Table II. All GDRs evaluated in this study, their clinical validity and additional data**.

**Supplementary Table III. PRISMA checklist for this study**

## References

Alhathal, N., Maddirevula, S., Coskun, S., Alali, H., Assoum, M., Morris, T., Deek, H. A., Hamed, S. A., Alsuhaibani, S., Mirdawi, A., Ewida, N., Al-Qahtani, M., Ibrahim, N., Abdulwahab, F., Altaweel, W., Dasouki, M. J., Assiri, A., Qabbaj, W., & Alkuraya, F. S. (2020). A genomics approach to male infertility. Genet Med, 22(12), 1967–1975. https://doi.org/10.1038/s41436-020-0916-0

Arafat, M., Harlev, A., Har-Vardi, I., Levitas, E., Priel, T., Gershoni, M., Searby, C., Sheffield, V. C., Lunenfeld, E., & Parvari, R. (2020). Mutation in CATIP (C2orf62) causes oligoteratoasthenozoospermia by affecting actin dynamics. J Med Genet. https://doi.org/10.1136/jmedgenet-2019-106825

Arango, N. A., Li, L., Dabir, D., Nicolau, F., Pieretti-Vanmarcke, R., Koehler, C., McCarrey, J. R., Lu, N., & Donahoe, P. K. (2013). Meiosis I arrest abnormalities lead to severe oligozoospermia in meiosis 1 arresting protein (M1ap)-deficient mice. Biol Reprod, 88(3), 76. https://doi.org/10.1095/biolreprod.111.098673

Bedoni, N., Haer-Wigman, L., Vaclavik, V., Tran, V. H., Farinelli, P., Balzano, S., Royer-Bertrand, B., El-Asrag, M. E., Bonny, O., Ikonomidis, C., Litzistorf, Y., Nikopoulos, K., Yioti, G. G., Stefaniotou, M. I., McKibbin, M., Booth, A. P., Ellingford, J. M., Black, G. C., Toomes, C., Inglehearn, C. F., Hoyng, C. B., Bax, N., Klaver, C. C., Thiadens, A. A., Murisier, F., Schorderet, D. F., Ali, M., Cremers, F. P., Andreasson, S., Munier, F. L., & Rivolta, C. (2016). Mutations in the polyglutamylase gene TTLL5, expressed in photoreceptor cells and spermatozoa, are associated with cone-rod degeneration and reduced male fertility. Hum Mol Genet, 25(20), 4546–4555. https://doi.org/10.1093/hmg/ddw282

Bourinaris, T., Smedley, D., Cipriani, V., Sheikh, I., Athanasiou-Fragkouli, A., Chinnery, P., Morris, H., Real, R., Harrison, V., Reid, E., Wood, N., Genomics England Research, C., Vandrovcova, J., Houlden, H., & Tucci, A. (2020). Identification of UBAP1 mutations in juvenile hereditary spastic paraplegia in the 100,000 Genomes Project. Eur J Hum Genet, 28(12), 1763–1768. https://doi.org/10.1038/s41431-020-00720-w

Camats, N., Fluck, C. E., & Audi, L. (2020). Oligogenic Origin of Differences of Sex Development in Humans. Int J Mol Sci, 21(5). https://doi.org/10.3390/ijms21051809

Cangiano, B., Swee, D. S., Quinton, R., & Bonomi, M. (2021). Genetics of congenital hypogonadotropic hypogonadism: peculiarities and phenotype of an oligogenic disease. Hum Genet, 140(1), 77–111. https://doi.org/10.1007/s00439-020-02147-1

Chen, S., Wang, G., Zheng, X., Ge, S., Dai, Y., Ping, P., Chen, X., Liu, G., Zhang, J., Yang, Y., Zhang, X., Zhong, A., Zhu, Y., Chu, Q., Huang, Y., Zhang, Y., Shen, C., Yuan, Y., Yuan, Q., Pei, X., Cheng, C. Y., & Sun, F. (2020). Whole-exome sequencing of a large Chinese azoospermia and severe oligospermia cohort identifies novel infertility causative variants and genes. Hum Mol Genet, 29(14), 2451–2459. https://doi.org/10.1093/hmg/ddaa101

Chiu, W., Hsun, Y. H., Chang, K. J., Yarmishyn, A. A., Hsiao, Y. J., Chien, Y., Chien, C. S., Ma, C., Yang, Y. P., Tsai, P. H., Chiou, S. H., Lin, T. Y., & Cheng, H. M. (2020). Current Genetic Survey and Potential Gene-Targeting Therapeutics for Neuromuscular Diseases. Int J Mol Sci, 21(24). https://doi.org/10.3390/ijms21249589

Colpi, G. M., Francavilla, S., Haidl, G., Link, K., Behre, H. M., Goulis, D. G., Krausz, C., & Giwercman, A. (2018). European Academy of Andrology guideline Management of oligo-astheno-teratozoospermia. Andrology, 6(4), 513–524. https://doi.org/10.1111/andr.12502

Cuvertino, S., Hartill, V., Colyer, A., Garner, T., Nair, N., Al-Gazali, L., Canham, N., Faundes, V., Flinter, F., Hertecant, J., Holder-Espinasse, M., Jackson, B., Lynch, S. A., Nadat, F., Narasimhan, V. M., Peckham, M., Sellers, R., Seri, M., Montanari, F., Southgate, L., Squeo, G. M., Trembath, R., van Heel, D., Venuto, S., Weisberg, D., Stals, K., Ellard, S., Genomics England Research, C., Barton, A., Kimber, S. J., Sheridan, E., Merla, G., Stevens, A., Johnson, C. A., & Banka, S. (2020). A restricted spectrum of missense KMT2D variants cause a multiple malformations disorder distinct from Kabuki syndrome. Genet Med, 22(5), 867–877. https://doi.org/10.1038/s41436-019-0743-3

de Mouzon, J., Chambers, G. M., Zegers-Hochschild, F., Mansour, R., Ishihara, O., Banker, M., Dyer, S., Kupka, M., & Adamson, G. D. (2020). International Committee for Monitoring Assisted Reproductive Technologies world report: assisted reproductive technology 2012dagger. Hum Reprod, 35(8), 1900–1913. https://doi.org/10.1093/humrep/deaa090

Domenice, S., Machado, A. Z., Ferreira, F. M., Ferraz-de-Souza, B., Lerario, A. M., Lin, L., Nishi, M. Y., Gomes, N. L., da Silva, T. E., Silva, R. B., Correa, R. V., Montenegro, L. R., Narciso, A., Costa, E. M., Achermann, J. C., & Mendonca, B. B. (2016). Wide spectrum of NR5A1-related phenotypes in 46,XY and 46,XX individuals. Birth Defects Res C Embryo Today, 108(4), 309–320. https://doi.org/10.1002/bdrc.21145

Ehrlich, S. (2015). Effect of fertility and infertility on longevity. Fertil Steril, 103(5), 1129–1135. https://doi.org/10.1016/j.fertnstert.2015.03.021

Eisenberg, M. L., Li, S., Behr, B., Cullen, M. R., Galusha, D., Lamb, D. J., & Lipshultz, L. I. (2014). Semen quality, infertility and mortality in the USA. Hum Reprod, 29(7), 1567–1574. https://doi.org/10.1093/humrep/deu106

Fagerberg, L., Hallstrom, B. M., Oksvold, P., Kampf, C., Djureinovic, D., Odeberg, J., Habuka, M., Tahmasebpoor, S., Danielsson, A., Edlund, K., Asplund, A., Sjostedt, E., Lundberg, E., Szigyarto, C. A., Skogs, M., Takanen, J. O., Berling, H., Tegel, H., Mulder, J., Nilsson, P., Schwenk, J. M., Lindskog, C., Danielsson, F., Mardinoglu, A., Sivertsson, A., von Feilitzen, K., Forsberg, M., Zwahlen, M., Olsson, I., Navani, S., Huss, M., Nielsen, J., Ponten, F., & Uhlen, M. (2014). Analysis of the human tissue-specific expression by genome-wide integration of transcriptomics and antibody-based proteomics. Mol Cell Proteomics, 13(2), 397–406. https://doi.org/10.1074/mcp.M113.035600

Fakhro, K. A., Elbardisi, H., Arafa, M., Robay, A., Rodriguez-Flores, J. L., Al-Shakaki, A., Syed, N., Mezey, J. G., Abi Khalil, C., Malek, J. A., Al-Ansari, A., Al Said, S., & Crystal, R. G. (2018). Point-of-care whole-exome sequencing of idiopathic male infertility. Genet Med, 20(11), 1365–1373. https://doi.org/10.1038/gim.2018.10

Goncalves, C. I., Fonseca, F., Borges, T., Cunha, F., & Lemos, M. C. (2017). Expanding the genetic spectrum of ANOS1 mutations in patients with congenital hypogonadotropic hypogonadism. Hum Reprod, 32(3), 704–711. https://doi.org/10.1093/humrep/dew354

Gou, L. T., Kang, J. Y., Dai, P., Wang, X., Li, F., Zhao, S., Zhang, M., Hua, M. M., Lu, Y., Zhu, Y., Li, Z., Chen, H., Wu, L. G., Li, D., Fu, X. D., Li, J., Shi, H. J., & Liu, M. F. (2017). Ubiquitination-Deficient Mutations in Human Piwi Cause Male Infertility by Impairing Histone-to-Protamine Exchange during Spermiogenesis. Cell, 169(6), 1090–1104 e1013. https://doi.org/10.1016/j.cell.2017.04.034

Jensen, T. K., Jacobsen, R., Christensen, K., Nielsen, N. C., & Bostofte, E. (2009). Good semen quality and life expectancy: a cohort study of 43,277 men. Am J Epidemiol, 170(5), 559–565. https://doi.org/10.1093/aje/kwp168

Kaplanis, J., Samocha, K. E., Wiel, L., Zhang, Z., Arvai, K. J., Eberhardt, R. Y., Gallone, G., Lelieveld, S. H., Martin, H. C., McRae, J. F., Short, P. J., Torene, R. I., de Boer, E., Danecek, P., Gardner, E. J., Huang, N., Lord, J., Martincorena, I., Pfundt, R., Reijnders, M. R. F., Yeung, A., Yntema, H. G., Deciphering Developmental Disorders, S., Vissers, L., Juusola, J., Wright, C. F., Brunner, H. G., Firth, H. V., FitzPatrick, D. R., Barrett, J. C., Hurles, M. E., Gilissen, C., & Retterer, K. (2020). Evidence for 28 genetic disorders discovered by combining healthcare and research data. Nature, 586(7831), 757–762. https://doi.org/10.1038/s41586-020-2832-5

Krausz, C., & Riera-Escamilla, A. (2018). Genetics of male infertility. Nat Rev Urol, 15(6), 369–384. https://doi.org/10.1038/s41585-018-0003-3

Leigh, M. W., Pittman, J. E., Carson, J. L., Ferkol, T. W., Dell, S. D., Davis, S. D., Knowles, M. R., & Zariwala, M. A. (2009). Clinical and genetic aspects of primary ciliary dyskinesia/Kartagener syndrome. Genet Med, 11(7), 473–487. https://doi.org/10.1097/GIM.0b013e3181a53562

Leroy, C., Fouveaut, C., Leclercq, S., Jacquemont, S., Boullay, H. D., Lespinasse, J., Delpech, M., Dupont, J. M., Hardelin, J. P., & Dode, C. (2008). Biallelic mutations in the prokineticin-2 gene in two sporadic cases of Kallmann syndrome. Eur J Hum Genet, 16(7), 865–868. https://doi.org/10.1038/ejhg.2008.15

Li, L., Gao, F., Fan, L., Su, C., Liang, X., & Gong, C. (2020). Disorders of Sex Development in Individuals Harbouring MAMLD1 Variants: WES and Interactome Evidence of Oligogenic Inheritance. Front Endocrinol (Lausanne), 11, 582516. https://doi.org/10.3389/fendo.2020.582516

Liu, C., Tu, C., Wang, L., Wu, H., Houston, B. J., Mastrorosa, F. K., Zhang, W., Shen, Y., Wang, J., Tian, S., Meng, L., Cong, J., Yang, S., Jiang, Y., Tang, S., Zeng, Y., Lv, M., Lin, G., Li, J., Saiyin, H., He, X., Jin, L., Toure, A., Ray, P. F., Veltman, J. A., Shi, Q., O’Bryan, M. K., Cao, Y., Tan, Y. Q., & Zhang, F. (2021). Deleterious variants in X-linked CFAP47 induce asthenoteratozoospermia and primary male infertility. Am J Hum Genet, 108(2), 309–323. https://doi.org/10.1016/j.ajhg.2021.01.002

Maione, L., Dwyer, A. A., Francou, B., Guiochon-Mantel, A., Binart, N., Bouligand, J., & Young, J. (2018). GENETICS IN ENDOCRINOLOGY: Genetic counseling for congenital hypogonadotropic hypogonadism and Kallmann syndrome: new challenges in the era of oligogenism and next-generation sequencing. Eur J Endocrinol, 178(3), R55–R80. https://doi.org/10.1530/EJE-17-0749

Markitantova, Y., & Simirskii, V. (2020). Inherited Eye Diseases with Retinal Manifestations through the Eyes of Homeobox Genes. Int J Mol Sci, 21(5). https://doi.org/10.3390/ijms21051602

Nagirnaja, L., Aston, K. I., & Conrad, D. F. (2018). Genetic intersection of male infertility and cancer. Fertil Steril, 109(1), 20–26. https://doi.org/10.1016/j.fertnstert.2017.10.028

Olesen, I. A., Andersson, A. M., Aksglaede, L., Skakkebaek, N. E., Rajpert-de Meyts, E., Joergensen, N., & Juul, A. (2017). Clinical, genetic, biochemical, and testicular biopsy findings among 1,213 men evaluated for infertility. Fertil Steril, 107(1), 74–82 e77. https://doi.org/10.1016/j.fertnstert.2016.09.015

Oud, M. S., Volozonoka, L., Friedrich, C., Kliesch, S., Nagirnaja, L., Gilissen, C., O’Bryan, M. K., McLachlan, R. I., Aston, K. I., Tuttelmann, F., Conrad, D. F., & Veltman, J. A. (2021). Lack of evidence for a role of PIWIL1 variants in human male infertility. Cell, 184(8), 1941–1942. https://doi.org/10.1016/j.cell.2021.03.001

Oud, M. S., Volozonoka, L., Smits, R. M., Vissers, L., Ramos, L., & Veltman, J. A. (2019). A systematic review and standardized clinical validity assessment of male infertility genes. Hum Reprod, 34(5), 932–941. https://doi.org/10.1093/humrep/dez022

Pan, J., Eckardt, S., Leu, N. A., Buffone, M. G., Zhou, J., Gerton, G. L., McLaughlin, K. J., & Wang, P. J. (2009). Inactivation of Nxf2 causes defects in male meiosis and age-dependent depletion of spermatogonia. Dev Biol, 330(1), 167–174. https://doi.org/10.1016/j.ydbio.2009.03.022

Pleuger, C., Lehti, M. S., Dunleavy, J. E., Fietz, D., & O’Bryan, M. K. (2020). Haploid male germ cells-the Grand Central Station of protein transport. Hum Reprod Update, 26(4), 474–500. https://doi.org/10.1093/humupd/dmaa004

Punab, M., Poolamets, O., Paju, P., Vihljajev, V., Pomm, K., Ladva, R., Korrovits, P., & Laan, M. (2017). Causes of male infertility: a 9-year prospective monocentre study on 1737 patients with reduced total sperm counts. Hum Reprod, 32(1), 18–31. https://doi.org/10.1093/humrep/dew284

Rehm, H. L. (2017). Evolving health care through personal genomics. Nat Rev Genet, 18(4), 259–267. https://doi.org/10.1038/nrg.2016.162

Schultz, N., Hamra, F. K., & Garbers, D. L. (2003). A multitude of genes expressed solely in meiotic or postmeiotic spermatogenic cells offers a myriad of contraceptive targets. Proc Natl Acad Sci U S A, 100(21), 12201–12206. https://doi.org/10.1073/pnas.1635054100

Sironen, A., Shoemark, A., Patel, M., Loebinger, M. R., & Mitchison, H. M. (2020). Sperm defects in primary ciliary dyskinesia and related causes of male infertility. Cell Mol Life Sci, 77(11), 2029–2048. https://doi.org/10.1007/s00018-019-03389-7

Smith, E. D., Radtke, K., Rossi, M., Shinde, D. N., Darabi, S., El-Khechen, D., Powis, Z., Helbig, K., Waller, K., Grange, D. K., Tang, S., & Farwell Hagman, K. wD. (2017). Classification of Genes: Standardized Clinical Validity Assessment of Gene-Disease Associations Aids Diagnostic Exome Analysis and Reclassifications. Hum Mutat, 38(5), 600–608. https://doi.org/10.1002/humu.23183

Toth, B., Baston-Bust, D. M., Behre, H. M., Bielfeld, A., Bohlmann, M., Buhling, K., Dittrich, R., Goeckenjan, M., Hancke, K., Kliesch, S., Kohn, F. M., Krussel, J., Kuon, R., Liebenthron, J., Nawroth, F., Nordhoff, V., Pinggera, G. M., Rogenhofer, N., Rudnik-Schoneborn, S., Schuppe, H. C., Schuring, A., Seifert-Klauss, V., Strowitzki, T., Tuttelmann, F., Vomstein, K., Wildt, L., Wischmann, T., Wunder, D., & Zschocke, J. (2019). Diagnosis and Treatment Before Assisted Reproductive Treatments. Guideline of the DGGG, OEGGG and SGGG (S2k Level, AWMF Register Number 015-085, February 2019) - Part 2, Hemostaseology, Andrology, Genetics and History of Malignant Disease. Geburtshilfe Frauenheilkd, 79(12), 1293–1308. https://doi.org/10.1055/a-1017-3478

Toure, A., Martinez, G., Kherraf, Z. E., Cazin, C., Beurois, J., Arnoult, C., Ray, P. F., & Coutton, C. (2021). The genetic architecture of morphological abnormalities of the sperm tail. Hum Genet, 140(1), 21–42. https://doi.org/10.1007/s00439-020-02113-x

Tu, C., Nie, H., Meng, L., Wang, W., Li, H., Yuan, S., Cheng, D., He, W., Liu, G., Du, J., Gong, F., Lu, G., Lin, G., Zhang, Q., & Tan, Y. Q. (2020). Novel mutations in SPEF2 causing different defects between flagella and cilia bridge: the phenotypic link between MMAF and PCD. Hum Genet, 139(2), 257–271. https://doi.org/10.1007/s00439-020-02110-0

Tuttelmann, F., Ruckert, C., & Ropke, A. (2018). Disorders of spermatogenesis: Perspectives for novel genetic diagnostics after 20 years of unchanged routine. Med Genet, 30(1), 12–20. https://doi.org/10.1007/s11825-018-0181-7

Uhlen, M., Fagerberg, L., Hallstrom, B. M., Lindskog, C., Oksvold, P., Mardinoglu, A., Sivertsson, A., Kampf, C., Sjostedt, E., Asplund, A., Olsson, I., Edlund, K., Lundberg, E., Navani, S., Szigyarto, C. A., Odeberg, J., Djureinovic, D., Takanen, J. O., Hober, S., Alm, T., Edqvist, P. H., Berling, H., Tegel, H., Mulder, J., Rockberg, J., Nilsson, P., Schwenk, J. M., Hamsten, M., von Feilitzen, K., Forsberg, M., Persson, L., Johansson, F., Zwahlen, M., von Heijne, G., Nielsen, J., & Ponten, F. (2015). Proteomics. Tissue-based map of the human proteome. Science, 347(6220), 1260419. https://doi.org/10.1126/science.1260419

Wang, X., Jin, H., Han, F., Cui, Y., Chen, J., Yang, C., Zhu, P., Wang, W., Jiao, G., Wang, W., Hao, C., & Gao, Z. (2017). Homozygous DNAH1 frameshift mutation causes multiple morphological anomalies of the sperm flagella in Chinese. Clin Genet, 91(2), 313–321. https://doi.org/10.1111/cge.12857

Whatley, M., Francis, A., Ng, Z. Y., Khoh, X. E., Atlas, M. D., Dilley, R. J., & Wong, E. Y. M. (2020). Usher Syndrome: Genetics and Molecular Links of Hearing Loss and Directions for Therapy. Front Genet, 11, 565216. https://doi.org/10.3389/fgene.2020.565216

Whitfield, M., Thomas, L., Bequignon, E., Schmitt, A., Stouvenel, L., Montantin, G., Tissier, S., Duquesnoy, P., Copin, B., Chantot, S., Dastot, F., Faucon, C., Barbotin, A. L., Loyens, A., Siffroi, J. P., Papon, J. F., Escudier, E., Amselem, S., Mitchell, V., Toure, A., & Legendre, M. (2019). Mutations in DNAH17, Encoding a Sperm-Specific Axonemal Outer Dynein Arm Heavy Chain, Cause Isolated Male Infertility Due to Asthenozoospermia. Am J Hum Genet, 105(1), 198–212. https://doi.org/10.1016/j.ajhg.2019.04.015

Wilfert, A. B., Chao, K. R., Kaushal, M., Jain, S., Zollner, S., Adams, D. R., & Conrad, D. F. (2016). Genome-wide significance testing of variation from single case exomes. Nat Genet, 48(12), 1455–1461. https://doi.org/10.1038/ng.3697

Winters, T., McNicoll, F., & Jessberger, R. (2014). Meiotic cohesin STAG3 is required for chromosome axis formation and sister chromatid cohesion. EMBO J, 33(11), 1256–1270. https://doi.org/10.1002/embj.201387330

Yang, F., De La Fuente, R., Leu, N. A., Baumann, C., McLaughlin, K. J., & Wang, P. J. (2006). Mouse SYCP2 is required for synaptonemal complex assembly and chromosomal synapsis during male meiosis. J Cell Biol, 173(4), 497–507. https://doi.org/10.1083/jcb.200603063

Yatsenko, A. N., Georgiadis, A. P., Ropke, A., Berman, A. J., Jaffe, T., Olszewska, M., Westernstroer, B., Sanfilippo, J., Kurpisz, M., Rajkovic, A., Yatsenko, S. A., Kliesch, S., Schlatt, S., & Tuttelmann, F. (2015). X-linked TEX11 mutations, meiotic arrest, and azoospermia in infertile men. N Engl J Med, 372(22), 2097–2107. https://doi.org/10.1056/NEJMoa1406192

Yuan, P., Yang, C., Ren, Y., Yan, J., Nie, Y., Yan, L., & Qiao, J. (2020). A novel homozygous mutation of phospholipase C zeta leading to defective human oocyte activation and fertilization failure. Hum Reprod, 35(4), 977–985. https://doi.org/10.1093/humrep/dez293

Zitzmann, M., Aksglaede, L., Corona, G., Isidori, A. M., Juul, A., T’Sjoen, G., Kliesch, S., D’Hauwers, K., Toppari, J., Slowikowska-Hilczer, J., Tuttelmann, F., & Ferlin, A. (2021). European academy of andrology guidelines on Klinefelter Syndrome Endorsing Organization: European Society of Endocrinology. Andrology, 9(1), 145–167. https://doi.org/10.1111/andr.12909

