## Supplementary Table III for "A systematic review of the validated monogenic causes of human male infertility: 2020 update and a discussion of emerging gene-disease relationships"

**Supplementary Table 1.** Search strategy, inclusion and exclusion criteria

| | |
|---|---|
| **Search term Pubmed** | "Infertility, Male/genetics"[Mesh]<br><br>OR<br><br>"Male sterility due to Y-chromosome deletions" [Supplementary Concept]<br><br>OR<br><br>(("genetics"[MeSH Terms] OR "genetics"[Subheading] OR "genetics"[All Fields] OR “genetic”[All Fields] OR "Genetic Variation"[Mesh] OR “variant”[All Fields] OR “variants”[All Fields] OR “CNV”[All Fields] OR “SNV”[All Fields] OR “SNP”[All Fields] OR “VUS”[All Fields] OR “mutation”[All Fields] OR “mutations”[All Fields] OR “polymorphism”[All Fields] OR “polymorphisms”[All Fields] OR "Karyotype"[Mesh] OR “Karyotype”[All fields] OR "Chromosomes, Human, Y/genetics"[Mesh] OR ((“Y chromosome”[All Fields] OR “chromosome Y”[All Fields] OR “Y-chromosome”[All Fields]) AND “microdeletion”[All Fields]) OR “AZFc deletion”[All Fields] OR “AZFb deletion”[All Fields] OR “AZFa deletion”[All Fields] OR “gr/gr deletion”[All Fields])<br><br>AND<br><br>("infertility, male"[MeSH Terms] OR ("infertility"[All Fields] AND "male"[All Fields]) OR "male infertility"[All Fields] OR ("male"[All Fields] AND "infertility"[All Fields]) OR “infertile men”[All Fields] OR "azoospermia"[MeSH Terms] OR "azoospermia"[All Fields] OR “Sertoli Cell-Only Syndrome”[MeSH] OR “Sertoli Cell-Only”[All Fields] OR “Aspermia”[All Fields]<br>OR<br>"oligospermia"[MeSH Terms] OR "oligospermia"[All Fields] OR "oligozoospermia"[All Fields] OR "cryptozoospermia"[All Fields] OR “hypospermatogenesis”[All Fields] OR “spermatogenic failure”[All Fields]<br>OR<br>"asthenozoospermia"[MeSH Terms] OR "asthenozoospermia"[All Fields] OR “Asthenospermia”[All Fields] OR “oligoasthenoteratozoospermia”[All Fields] OR "multiple morphological abnormalities of the sperm flagella "[All Fields] OR "MMAF"[All Fields] OR “Dysplasia of the fibrous sheath”[All Fields] OR "Kartagener Syndrome"[Mesh] OR "Kartagener Syndrome"[All Fields] OR "Primary ciliary dyskinesia"[All Fields] OR "teratozoospermia"[MeSH Terms] OR "teratozoospermia"[All Fields] OR “teratospermia”[All Fields] OR "globozoospermia"[All Fields] OR "macrozoospermia"[All Fields] OR ("acephalic"[All Fields] AND ("spermatozoa"[MeSH Terms] OR "spermatozoa"[All Fields])) OR "Congenital bilateral aplasia of vas deferens" [Supplementary Concept] OR “Congenital bilateral aplasia of vas deferens”[All Fields] OR “Congenital bilateral absence of vas deferens”[All Fields] OR “CBAVD”[All Fields] OR "Vas Deferens/abnormalities"[Mesh] OR "Cryptorchidism"[Mesh] OR "Cryptorchidism"[All Fields] OR "Persistent Mullerian duct syndrome"[Supplementary Concept] OR "Persistent Mullerian duct syndrome"[All Fields] OR "Hypospadias"[Mesh] OR "Hypospadias"[All Fields]<br>OR |

| | |
|---|---|
| | "Kallmann Syndrome"[Mesh] OR "Kallmann Syndrome"[All fields] OR "Hypogonadism"[Mesh] OR "Hypogonadism"[All fields] OR "Klinefelter Syndrome"[Mesh] OR "Klinefelter Syndrome"[All fields] OR "XXY"[All fields] OR "46, XY Disorders of Sex Development"[Mesh] OR "46, XX Disorders of Sex Development"[Mesh] OR "46, XY Disorders of Sex Development"[All Fields] OR "46,XY Disorders of Sex Development"[All Fields] OR "46, XX Disorders of Sex Development"[All Fields] OR "46,XX Disorders of Sex Development"[All Fields] OR "46, XY Disorder of Sex Development"[All Fields] OR "46,XY Disorder of Sex Development"[All Fields] OR "46, XX Disorder of Sex Development"[All Fields] OR "46,XX Disorder of Sex Development"[All Fields] OR "Androgen-Insensitivity Syndrome"[Mesh] OR "androgen insensitivity syndrome"[All fields] OR "Leydig Cell Hypoplasia" [Supplementary Concept] OR "Receptors, LH"[Mesh] OR "Leydig cell hypoplasia"[All Fields] OR (("Gonadal Dysgenesis/genetics"[Mesh] OR "Gonadal Dysgenesis, 46,XY"[Mesh] OR "Gonadal Dysgenesis" [All Fields]) AND ("46,XY"[All Fields] OR "46, XY"[All Fields] OR "46XY"[All Fields]))) <br><br> NOT <br><br> ("Plant Infertility"[Mesh] OR "Fungi"[Mesh] OR "Cattle"[Mesh] OR "Swine"[Mesh] OR "Intellectual Disability"[Mesh] OR ("Heart Defects, Congenital"[Mesh] NOT ("Dextrocardia"[Mesh] OR "Kartagener Syndrome"[Mesh] OR "Noonan Syndrome"[Mesh] OR "Turner Syndrome"[Mesh] OR "Heterotaxy syndrome"[Mesh])) OR "Leukoencephalopathies"[Mesh] OR "Preimplantation Diagnosis"[Mesh] OR "Prenatal Diagnosis"[Mesh] OR "Leukemia"[Mesh] OR "Histocompatibility Antigens Class I"[Mesh] OR "Histocompatibility Antigens Class II"[Mesh])) <br><br> AND <br><br> 1900:2020/03/11[dp] |
| **Inclusion criteria** | 1. Publish in peer-reviewed journal<br>2. Studies concentrating on male infertility and/or defective genitourinary development<br>3. Studies conducted in human patients<br>4. Studies concentrating on finding a genetic cause<br>5. Studies presenting a monogenic form of infertility<br>6. Studies presenting original data |
| **Exclusion criteria title/abstract screening** | 1. Publication not in English<br>2. Studies investigating animals/plants/fungi/prokaryotes<br>3. Study topic is irrelevant and/or includes one of the following examples:<br>- Female infertility<br>- Studies investigating polygenic or multifactorial forms of infertility<br>- Sperm aneuploidies or chromosomal anomalies without mentioning constitutional chromosomal anomalies<br>- Sperm DNA fragmentation or damage<br>- mtDNA copy load<br>- Variants described in population databases<br>- Epigenetics/methylome<br>- Sperm mRNA expression<br>- Studies about patients with Prader scale 0-3 (included: scale 4, 5 and 6) |

| | |
|---|---|
|  | - Syndromes characterized by severe physical or intellectual disability including CHARGE, Werner, Noonan, 4H, Opitz G/BBB, Gordon Holmes, Nijmegen breakage, Cabezas, MEHMO, Dilated Cardiomyopathy and Ataxia, Boucher-Neuhauser, Hartsfield syndrome, Aarskog-Scott, Huppke-Brendel, Brooks, Juberg-Marsidi, Warburg Micro, Prune Belly<br>- Other syndromes: Thalassemia, Myhre, Waardenburg, Bardet-Biedl, Wilms Tumor, Alström, Complete Androgen Insensitivity syndrome, Swyer syndrome, Frasier syndrome, Woodhouse-Sakati, Cystic Fibrosis, Cushing's syndrome, Gorlin-Goltz |
| **Exclusion criteria full text screening** | 4. Studies investigating genetic risk factors or associations without presuming a direct consequence of the variant on the gene/protein function<br>5. Deletion of multiple genes in AZF regions<br>6. Deletion or duplication of multiple genes<br>7. Studies presenting chromosomal aneuploidies or rearrangements without charging a (set of)affected gene(s)<br>8. Full text reveals study topic is irrelevant (based on exclusion criterion 3)<br>9. Full text not available |

| | |
|---|---|
| Web of Science search term | TS=("male infertility" or "infertile men" or "azoospermia" or "sertoli cell only" or "aspermia" or "oligospermia" or "oligozoospermia" or "cryptozoospermia" or “hypospermatogenesis” or “spermatogenic failure” or "asthenozoospermia" or “Asthenospermia” or “oligoasthenoteratozoospermia” or "multiple morphological abnormalities of the sperm flagella" or "MMAF" or “Dysplasia of the fibrous sheath” or "Kartagener Syndrome" or "Primary ciliary dyskinesia" or "teratozoospermia" or “teratospermia” or "globozoospermia" or "macrozoospermia" or "acephalic spermatozoa" or "acephalic sperm" or "Congenital bilateral aplasia of vas deferens" or "CBAVD" or "Cryptorchidism" or "Persistent Mullerian duct syndrome" or "Hypospadias" or "Kallmann Syndrome" or "Hypogonadism" or "Klinefelter Syndrome" or “XXY” or "46, XY Disorders of Sex Development" or "46,XY Disorders of Sex Development" or "46, XX Disorders of Sex Development" or "46,XX Disorders of Sex Development" or "46, XY Disorder of Sex Development" or "46,XY Disorder of Sex Development" or "46, XX Disorder of Sex Development" or "46,XX Disorder of Sex Development" or "Androgen-Insensitivity Syndrome" or “androgen insensitivity syndrome” or "Leydig Cell Hypoplasia" or "Gonadal Dysgenesis") NOT PMID=(1* or 2* or 3* or 4* or 5* or 6* or 7* or 8* or 9* or 0*)<br>AND<br>TS=("genetics" or "genetic" or "genetic variation" or “variant” or “variants” or “VUS” or “CNV” or “SNV” or “SNP” or "mutation" or "mutations" or "polymorphism" or "polymorphisms" or "karyotype" or "AZFc deletion" or "AZFa deletion" or "AZFb deletion" or "gr/gr deletion" or "chromosome" or "Y-chromosome")<br>AND<br>PY=(1900-2020)<br>Refined by: [excluding] DOCUMENT TYPES: ( MEETING ABSTRACT ) |
| | PMIDs_clean <-<br>str_remove_all(PMIDs$V1,"\n")<br>cat(PMIDs_clean, sep=" or ") |
